# COVID-19 in persons affected by Hansen’s disease in Brazil

**DOI:** 10.1101/2020.12.11.20247262

**Authors:** Patrícia D. Deps, Taynah A. R. Repsold, Claudio G. Salgado, Raquel de Carvalho Bouth, Selma Regina Penha Silva Cerqueira, Marisa Simon Brezinscki, Rebeca Ruppert Galarda Baptista Peixoto, Jaison Antonio Barreto, Andrea M. A. Fonseca, Marlene L. S. Peixoto, Seyna Ueno R. Mendes, Rafael Pereira Rabelo Mendes, Pedro Paulo dos Santos Oliveira, Ciro Martins Gomes

## Abstract

**Background:** Hansen’s disease (HD) is endemic in Brazil, a country with the third highest number of COVID-19 cases in the world and the second highest number of COVID-19 deaths. COVID-19 in persons affected by HD has not been described at population level in this country.

**Methods:** We collated numbers of COVID-19 cases and deaths among patients who were receiving routine treatment for HD at six centres across Brazil (Belém, Bauru, Brasília, Vitória, Petrolina, Palmas) between 1^st^ March and 10^th^ December 2020.

**Results:** Of 1,333 HD patients receiving treatment, 70 (5.2%) reported having had COVID-19. Almost all patients (97% (1,296/1,333)) including all but one of the COVID-19 cases were receiving MDT comprising rifampicin (600mg once per month), dapsone (100mg daily), and clofazimine (50 mg daily plus 300 mg once per month). Four patients died, including a patient in their 30’s on MDT who had a severe type 2 HD reaction (erythema nodosum leprosum) and who was taking clofazimine 100mg daily.

**Conclusions:** We cannot determine from these preliminary data whether persons affected by Hansen’s disease have a higher or lower risk of COVID-19 and related mortality compared with the general population. We will continue to monitor the effects of COVID-19 in persons affected by and treated for HD and extend this to monitor SARS-CoV-2 vaccine effectiveness in this group of patients.

## Introduction

Hansen’s disease remains endemic in Brazil, with 30,000 new cases diagnosed each year [1]. Brazil is also one of the countries most severely impacted by the COVID-19 pandemic, reporting 6 million cases and 170,000 deaths to the end of November 2020 [2]. As a first step to investigating occurrence and outcomes of COVID-19 in persons affected by Hansen’s disease, we analysed data from six centres in Brazil treating patients with Hansen’s disease from 1^st^ March to 10^th^ December 2020. Routine multidrug therapy for HD includes clofazimine, which was demonstrated to have SARS-CoV-2 antiviral activity in an animal model. This has led to the suggestion that clofazimine might have effects against COVID-19 in humans. We review the history of clofazimine as a treatment for HD and discuss whether it merits investigation as a treatment for COVID-19.

## Methods

Six HD treatment centres were recruited to the study, one each in the municipalities of Belém (state of Pará), Bauru (São Paulo), Brasília (Goias), Vitória (Espírito Santo), Petrolina (Pernambuco), and Palmas (Tocantins). Numbers of patients treated for HD at each centre were collated together with reported diagnoses of COVID-19 and deaths attributed to COVID-19 (from clinic records). Patients attending one centre (Brasília) were routinely tested (by PCR) for COVID-19. Background cumulative incidence of COVID-19 in each municipality was obtained from publicly available national surveillance data (https://covid.saude.gov.br/). Ethical approval was obtained from the research ethics committee (Comitê de Ética em Pesquisa, CEP) of the Universidade Federal do Espírito Santo (UFES), certificate no. 40347520.5.0000.5060.

## Results

Of 1,333 patients attending the six clinics between March 1^st^ and December 10^th^, 2020, 70 (5.2%) reported having had COVID-19 (**Table 1**). Most of the patients (97% (1,296/1,333)) including all but one of the COVID-19 cases were receiving MDT comprising rifampicin (600mg once per month), dapsone (100mg daily), and clofazimine (50 mg daily plus 300 mg once per month). Four patients died, including a patient in their 30’s on MDT who had a severe type 2 Hansen’s disease reaction (erythema nodosum leprosum, ENL) and who was taking clofazimine 100mg daily.

**Table 1.** **Persons affected by Hansen’s disease and COVID-19 diagnoses at four centres in Brazil, from March 1**^**st**^ **to December 10**^**th**^, **2020**

| Centre | Persons with Hansen's disease | Persons diagnosed with COVID-19 | COVID-19 deaths | Background COVID-19 cumulative incidence<sup>‡</sup> |
| --- | --- | --- | --- | --- |
| Belém | 83 | 6 (7.2%) | 0 | 3.5% |
| Bauru | 100 | 1 (1.0%) | 0 | 4.4% |
| Brasília | 98 | 13 (13.3%)<sup>†</sup> | 0 | 7.8% |
| Vitória | 66 | 1 (1.5%) | 1 | 6.9% |
| Petrolina | 463 | 14 (3.0%) | 1 | 2.7% |
| Palmas | 523 | 35 (6.7%) | 2 | 6.6% |
| Total | 1333 | 70 (5.2%) | 4 | 6.1% |

<sup>†</sup> Hansen's disease patients at this centre were tested routinely (PCR) for COVID-19
<sup>‡</sup> National surveillance data to December 10<sup>th</sup>, 2020, from https://covid.saude.gov.br/

## Discussion

This is the first population-level report of cases of COVID-19 among persons affected by HD. Whilst the cumulative incidence of COVID-19 in patients receiving treatment for HD appears similar to the background cumulative incidence of COVID-19 in each municipality, we cannot determine from these preliminary data whether persons affected by Hansen’s disease have a higher or lower risk of COVID-19 and associated mortality than the general population. This comparison would require adjustment for differences in age, sex and other factors. The relatively high rate of COVID-19 among HD patients in Brasília reflects routine testing for COVID-19 at this centre, a practice that other centres might consider adopting subject to availability of resources. We will continue to monitor the effects of COVID-19 in persons affected by and treated for HD and we plan to extend this to monitor SARS-CoV-2 vaccine effectiveness in this group of patients.

Both COVID-19 and reactional states in Hansen’s disease can present a severe cytokine storm, with markedly higher levels of pro-inflammatory cytokines including interferons, tumour necrosis factors, interleukins, and chemokines [3, 4]. In relation to the patient in our preliminary data who had ENL and who died from COVID-19, both conditions share immunological aspects related to generalized neutrophil infiltration: in SARS-CoV-2 infection, extensive neutrophil infiltration in pulmonary capillaries [5]; in ENL, extensive dermal neutrophil infiltration [6]. COVID-19 neutrophils may trigger ENL or increase its severity, or *vice-versa*. Systemic corticosteroids are important drugs for the treatment of COVID-19 and Hansen’s disease reactions. Prolonged, high-dose treatment with prednisolone (for 12-14 weeks) causing immunosuppression would increase risks associated with SARS-CoV-2 infection [7].

A recent large-scale drug repurposing study identified clofazimine (N,5-bis(4-chlorophenyl)-3-(1-methylethylimino)-5H-phenazin-2-amine) as a potential SARS-CoV-2 antiviral agent [8]. In a follow-up study, clofazimine *in vitro* and in hamsters demonstrated inhibitory activity against SARS-CoV-2 replication and reduced expression of IL-6, TNF-α, and CCR4 that suggested potential to reduce the cytokine storm that can occur in COVID-19 [9]. The possible effects of clofazimine on severity of SARS-CoV-2 infection have not been studied in people, but a phase II clinical trial was registered (Hong Kong, China) in July 2020 [<u>NCT04465695</u>].

Other than off-label use for dermatological conditions and atypical mycobacterial infections [10], clofazimine is used routinely only for the treatment of Hansen’s disease as a component of the WHO multidrug therapy (MDT) regimen comprising dapsone, clofazimine and rifampicin [11]. Clofazimine is a riminophenazine dye with bacteriostatic and anti-inflammatory actions, the mechanisms of which are largely unknown but possibly including direct activity affecting bacterial DNA [10]. Clofazimine is well tolerated in the current dosage for Hansen’s disease (50mg daily plus 300mg monthly), with mild side effects including dryness and hyperpigmentation of the skin [12], ocular conjunctiva and organic fluids [13]. At doses above 100mg daily, clofazimine can cause gastrointestinal symptoms; these can manifest as small bowel syndrome, a more serious side effect characterized by persistent diarrhoea, weight loss and abdominal pain [13-15]. The skin discoloration due to medium to long-term use of clofazimine can cause distress and has been associated with the stigmatisation of persons affected by Hansen’s disease [16].

According to Yuan *et al*., the dose of clofazimine which demonstrated efficacy against SARS-CoV-2 in hamsters was 25 mg/kg, corresponding to 200 mg/day in humans [9]. Reports from the 1970’s described use of clofazimine at 100, 200 and 300 mg/day for more than two months with few serious side effects [15, 17, 18]. Opromolla *et al*. (1972) reported excellent tolerance and good therapeutic results using 300 mg/day, and clofazimine was considered to have potent anti-inflammatory activity that translated into neutralisation of or prophylaxis against Hansen’s disease reactions [15]. Doses of 300mg/day are still included in national guidelines for the control of reactional states as an alternative to systemic corticosteroids in immunosuppressive doses [19].

Silva *et al*. (1972) commented that clofazimine is easily absorbed through the oral route, does not reach high concentrations in the blood because it is quickly captured by the cells of the endothelial reticulum system, and is deposited preferentially in the lungs, liver and skin; it is eliminated in urine, to which it lends a reddish colour, and by the sebaceous glands; dosages in excess of 400 mg/day were not recommended because of gastrointestinal side effects [18]. Clofazimine appears not to accumulate in blood or tissues, therefore in cases of treatment interruption a therapeutic effect is not guaranteed [20]. Conversely, recent pharmacokinetic studies in mouse models point to a possible cumulative effect of clofazimine after 2 weeks to 2 months of regular use and a possible increase in its half-life [21].

For as long as COVID-19 has few prophylactic drugs, and whilst awaiting distribution of vaccines which may take time to reach populations in remote and less well-resourced areas, it has been suggested that clofazimine merits further investigation in relation to possible protective effects against COVID-19. Considering the acute evolution time and high case fatality of COVID-19, side effects from higher doses or prolonged treatment would not be a limitation if clofazimine proved to be effective in suppressing the virus. However, we have some doubts that data from this patient group can provide evidence regarding the potential effectiveness of clofazimine against COVID-19:

1. In most centres, occurrence of COVID-19 is reported by patients or their family members to the health care professionals at monthly consultations
2. Many Hansen’s disease patients have had reduced access to health care services during the COVID-19 pandemic
3. Shortages of MDT mean that treatment is not available in many centres or patients have stopped taking MDT during the COVID-19 pandemic
4. Effects of clofazimine can be investigated by comparing Hansen’s disease patients who are receiving or not receiving clofazimine, but this restricts the generalizability of findings to the general population and results might be confounded by indication
5. Effects of clofazimine can be investigated by comparing Hansen’s disease patients who are receiving clofazimine with the general population, but any results will be heavily confounded by pathophysiological effects of Hansen’s disease and differences between patients and the general population and the effect of clofazimine cannot be isolated from the other components of MDT
6. The dosage of clofazimine in MDT for Hansen’s disease (50 mg/day) is far below the equivalent dose (200 mg/day) extrapolated from Yuan *et al*. [9]

Although clofazimine has few side effects and is rarely replaced, antibiotics (including minocycline, ofloxacin, doxycycline, clarithromycin, moxifloxacin) are indicated to replace MDT components in cases where side effects are sufficiently serious [14, 22], or in cases of treatment failure and resistance. Corticosteroids and thalidomide are recommended to control Hansen’s disease reactions, and vitamin D, bisphosphonates, and aspirin to reduce treatment-related damage [23]. Analgesics, nonsteroidal anti-inflammatories, antibiotics, metformin, proton-pump inhibitors, amitriptyline, gabapentin, simvastatin are also used during treatment of Hansen’s disease [23]. This polypharmacy would further confound any estimated effects of clofazimine.

One clinical trial of oral clofazimine for COVID-19 has been registered, using oral clofazimine 100mg twice daily on day 1, then 100mg daily for 2 days, compared with the same regimen plus a 3-day course of interferon β-1b [<u>NCT04465695</u>]. To investigate the effectiveness of clofazimine against severity of SARS-CoV-2 infection, we think that a higher dose of clofazimine can be given safely for a longer period. We would propose a clinical trial including oral clofazimine 200 mg/day for around two weeks. The experience of Brazilian physicians in routine use of clofazimine, including in high doses for HD reactions, and the currently high incidence of COVID-19 would make Brazil an ideal setting for such a study. In the meantime, we will continue to monitor the effects of COVID-19 in persons affected by and treated for Hansen’s disease, and extend this to monitor SARS-CoV-2 vaccine efficacy in this group of patients.

## Data Availability

Data are available by request to the corresponding author.

